# Establishing a blood-stage controlled human malaria infection model with *Plasmodium falciparum* in healthy semi-immune Tanzanian adults

**DOI:** 10.64898/2026.10.25.26366323

**Authors:** Sarah E. Silk, Jo Salkeld, Omary Hassan, Neema Balige, Latipha Mohamed, Florence Milando, Jordan R. Barrett, Saumu Ahmed, Aina-Ekisha Kahatano, Enock Kessy, Prisca Kweyamba, Doris Quinkert, Omary Juma, Bakari Mwalimu, Mohamed Mohamed, Natalie G. Marchevsky, Lorenz M. Hofer, Mgeni Mohamed, Brian B. Tarimo, Hania Msami, Tunu G. Mwamlima, Wilmina F. Kalinga, Ibrahim A. Sasamalo, Ivanny M. Mtaka, Nasoro S. Lilolime, Francesca R. Donnellan, Martino Bardelli, Ababacar Diouf, Alison M. Lawrie, Rachel Roberts, Fay L. Nugent, Carole A. Long, Sarah Mswata, Mabula Kasubi, Jee-Sun Cho, Carolyn M. Nielsen, Simon J. Draper, Angela M. Minassian, Ally I. Olotu

**Affiliations:** Department of Paediatrics, Department of Biochemistry and Kavli Institute for Nanoscience Discovery, University of Oxford, Oxford, United Kingdom; Interventions and Clinical Trials Department, Ifakara Health Institute, Bagamoyo, Tanzania; Oxford Vaccine Group, Department of Paediatrics, University of Oxford, Oxford, United Kingdom; The National Institute for Health and Care Research (NIHR) Oxford Biomedical Research Centre, Oxford, United Kingdom; Swiss Tropical and Public Health Institute, Department of Research, Allschwil, Switzerland; Department of Medical Entomology and Parasitology, School of Public Health and Social Sciences, Muhimbili University of Health and Allied Sciences, Dar es Salaam, Tanzania; Department of Vector Biology, Liverpool School of Tropical Medicine, Liverpool, United Kingdom; Laboratory of Malaria and Vector Research, National Institute of Allergy and Infectious Diseases, National Institutes of Health, Rockville, MD, USA; The Jenner Institute, Nuffield Department of Medicine, University of Oxford, Oxford, United Kingdom

## Abstract

Controlled human malaria infection (CHMI) studies are a critical tool for the assessment and down-selection of potential malaria interventions, allowing detailed characterisation and understanding of host-parasite responses to inform both malaria vaccine and antimalarial drug development in humans. Blood-stage CHMI offers a specific means for proof-of-concept testing of blood-stage malaria vaccines, bypassing the pre-erythrocytic stages of malaria infection. Here, we establish a blood-stage CHMI model in an endemic region in a clinical study involving 12 healthy semi-immune male adult participants residing in Bagamoyo district, Tanzania. Participants were enrolled into relatively “low” (n=6) and “high” (n=6) malaria pre-exposure groups as defined by serum anti-schizont IgG levels. Participants were challenged with approximately 1000 erythrocytes infected with a chloroquine-sensitive Plasmodium falciparum 3D7 clone and closely monitored for 28 days at the Ifakara Health Institute clinical trial facility in Bagamoyo. Blood samples were collected for safety and immunology assessments, with venous blood samples for P. falciparum quantitative polymerase chain reaction (qPCR) taken once on day after challenge (C+1) and twice daily from two to 28 days after challenge (C+2 to C+28). Participants were treated once pre-defined diagnostic criteria were met or at day 28 (if criteria not met). The challenge inoculum was found to be safe with the severity grading of adverse events (AEs) in keeping with previous challenge studies. Overall, fewer AEs were reported as compared to malaria-naïve UK adults previously challenged with the same inoculum dose, despite a 50-fold higher treatment threshold being used in Tanzanian adults. Nine of 11 participants developed parasitaemia detectable by qPCR and one participant was retrospectively excluded from parasitaemia endpoint analysis due to non-3D7 type infection. The two participants who remained qPCR negative were both in the “high” pre-exposure group. Higher anti-schizont baseline serology status was associated with later median time to parasitaemia detection and time to pre-defined treatment criteria. These data demonstrate that blood-stage CHMI is safe and feasible in semi-immune adults in a malaria-endemic setting.

## INTRODUCTION

Controlled human malaria infection (CHMI) is an established and widely used tool to assess the efficacy of novel malaria candidate vaccines and antimalarial drugs. It has supported the investigation of host-parasite interactions [1–6], immune responses to the malaria parasite [7, 8], and identification of potential correlates of protection [9–11]. Malaria infection can be initiated by the bite of infected mosquitoes, administration of cryopreserved sporozoites via various routes, or intravenous administration of infected erythrocytes (iRBC) [5, 12–16]. Following infection, participants are closely monitored for blood-stage parasitaemia and diagnosed once a pre-defined parasitaemia threshold, or a pre-defined combination of malaria symptoms and parasitaemia threshold, is reached. Time to detectable parasitaemia, time to diagnosis, measures of the liver to blood inoculum (LBI), blood-stage parasite multiplication rate (PMR) or parasite reduction ratio are all outcome metrics that can be used to evaluate the efficacy of candidate vaccines or antimalarial drugs [6, 12, 15, 17, 18]. Additionally, assessment of other parameters such as immune biomarkers and genetic signatures in immune cells provide insight into malaria-specific immune responses and potential correlates of protection [19, 20].

Blood-stage CHMI bypasses the pre-erythrocytic stages of malaria infection but enables greater confidence in modelling of blood-stage parasite growth dynamics and thus offers improved power to observe partial efficacy of blood-stage vaccine candidates [9, 12, 21]. This is because blood-stage CHMI allows inoculation with a known and comparable number of iRBC across participants in a study, rather than a more variable LBI as inevitably occurs after mosquito-bite initiated sporozoite CHMI [12, 17].

CHMI studies initiated by inoculation with aseptic, purified, cryopreserved *P. falciparum* sporozoites (PfSPZ) have been undertaken in six African countries to-date [22]. However, blood-stage CHMI studies have not previously been reported in populations living in malaria-endemic regions. Although blood-stage CHMI studies in malaria-naïve populations have provided significant contributions to the development of malaria interventions and understanding of malaria immunity, and have established the safety of the model, transfer to malaria-endemic populations is now essential. Importantly, undertaking such studies in Africa will enable data on malaria interventions to be generated in target populations, including assessment of the impact of variation in population genetic background and of naturally-acquired immunity (NAI) to malaria arising from prior exposure [23, 24]. There is, therefore, a clear rationale to translate and establish the blood-stage CHMI model in malaria-endemic settings.

Here we report the results of a pilot blood-stage CHMI study in 12 semi-immune, healthy Tanzanian adult participants. We sought to assess the safety and feasibility of *P. falciparum* blood-stage CHMI in a malaria-endemic population and establish the blood-stage CHMI model in Tanzania for the first time.

## RESULTS

### Participants

Seventy-one individuals underwent screening, of which 59 were excluded. Fifteen participants met the eligibility criteria for the VAC083 trial and 12 were enrolled across two study Groups (relatively “high” malaria pre-exposure, n=6 [Group 1]; versus “low” malaria pre-exposure, n=6 [Group 2]) (**Figure 1**, **Supplementary Table 2**).

**Figure 1.**
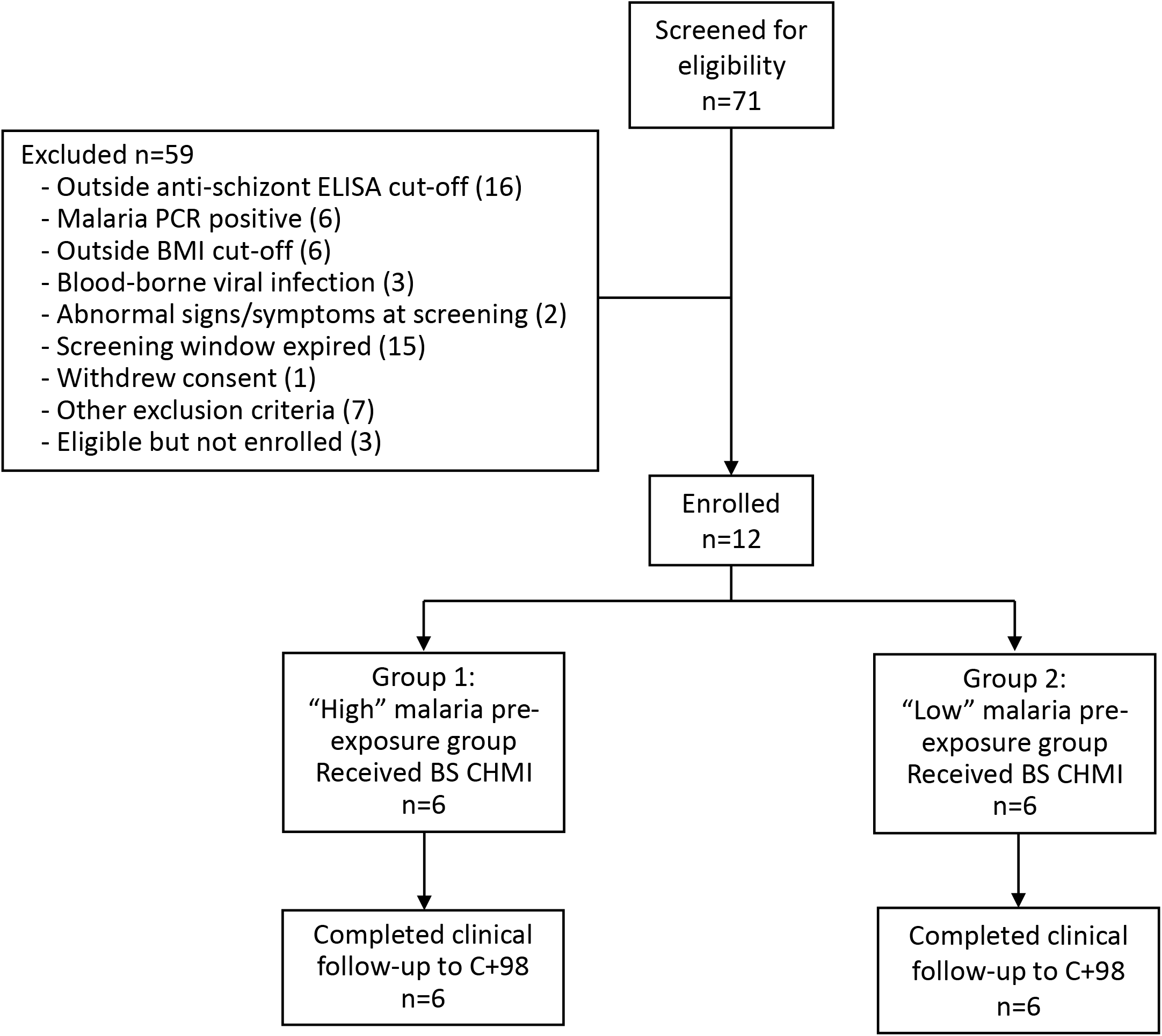
VAC083 trial profile. Seventy-one volunteers were screened, with 59 excluded and 12 participants enrolled into either relatively “high” or “low” malaria pre-exposure groups based on anti-schizont serum IgG ELISA. All participants underwent simultaneous 3D7 *P. falciparum* blood-stage controlled human malaria infection (BS CHMI) on 27^th^ July 2022. All participants completed follow-up until 98 days post-CHMI (C+98).

Demographics of the participants alongside baseline anti-schizont IgG antibody titre values are presented in **Table 1**. Median age for the high and low malaria pre-exposure groups was 22.6 years and 25.1 years, respectively. All participants enrolled in the study were male. The median anti-schizont IgG levels at baseline for the high and low pre-exposure groups were 2449 AU and 189 AU, respectively, i.e. ∼13-fold higher baseline anti-malaria schizont serum IgG responses in the relatively high pre-exposure group as compared to the low pre-exposure group; also group ranges did not overlap (**Figure 3A**). All twelve enrolled participants were inoculated on the 27^th^ July 2022 and completed all follow-up visits out to 98 days post-CHMI (C+98).

**Figure 2.**
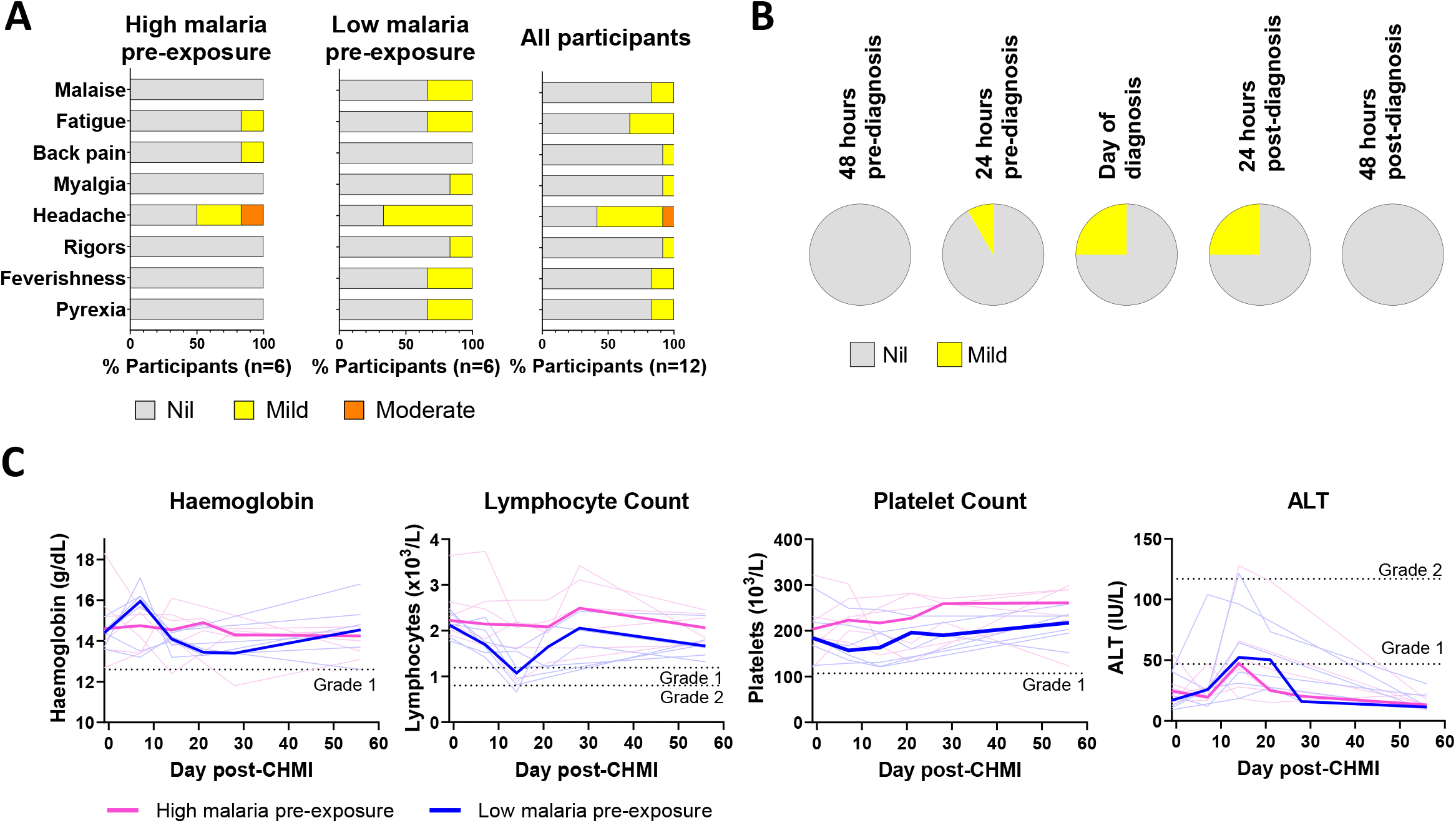
Adverse event analysis of 3D7 *P. falciparum* CHMI in participants with high and low malaria pre-exposure. (**A**) CMHI-related solicited adverse events (AEs) were recorded from day of CHMI until completion of treatment. The percentage of participants reporting each AE in each Group and across all participants is shown, with the maximum severity of each AE in each participant recorded. (**B**) The proportion of all participants reporting any solicited AE at each grade at each timepoint from 48 hours pre-diagnosis until 48 hours post-diagnosis is shown. Solicited AEs reported outside of this timeframe are not shown. (**C**) Haemoglobin, lymphocyte count, platelet count and alanine aminotransferase (ALT) levels are shown for each participant at C-1, C+7, C+14, C+21, C+28 and C+56 days in the high (pink) and low (blue) malaria pre-exposure groups. The median result for each group at each timepoint is shown in bold.

**Figure 3.**
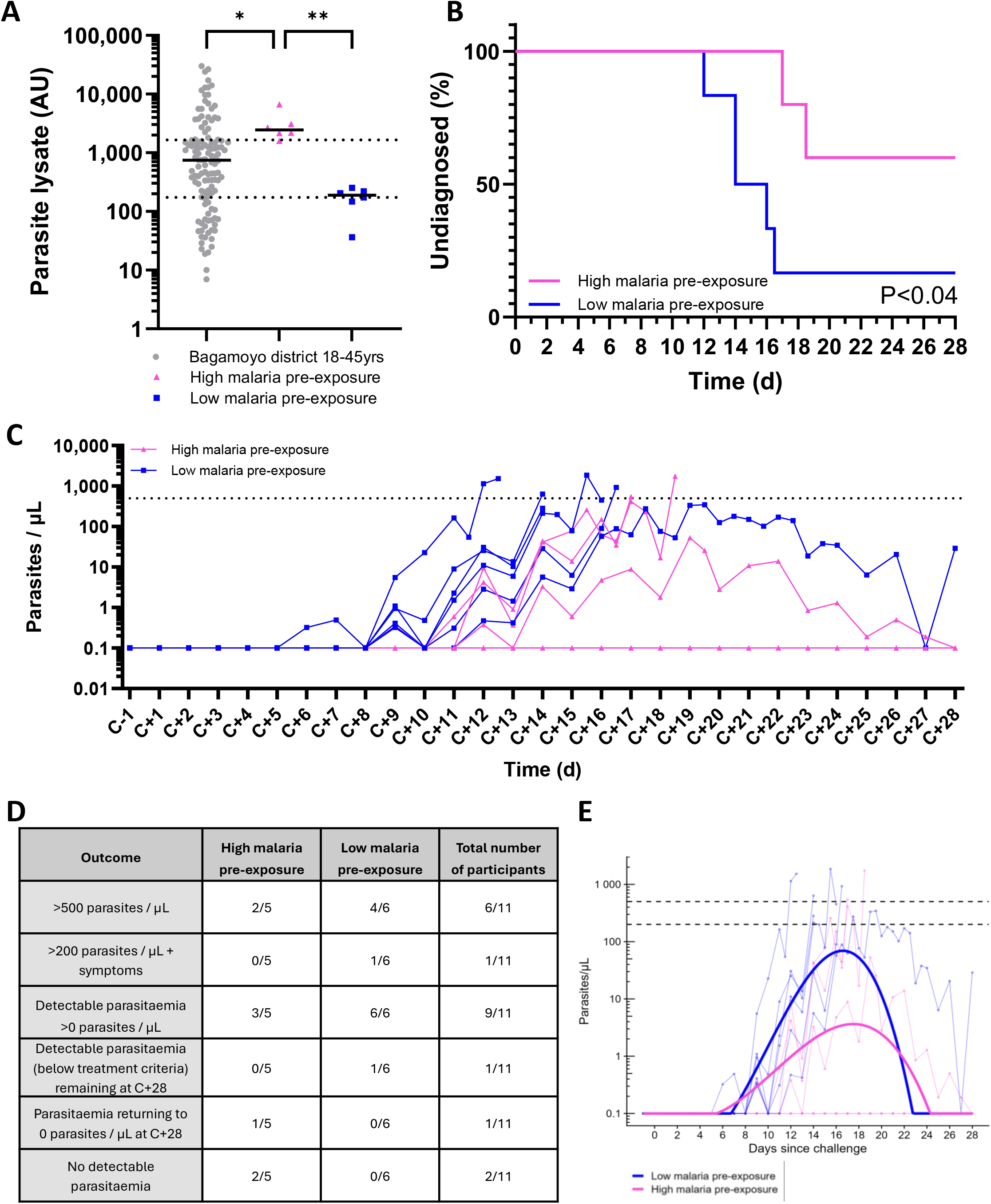
*P. falciparum* parasitaemia outcomes following blood-stage CHMI. (**A**) Median and individual baseline anti-parasite lysate total IgG responses for high (n=6) and low (n=6) malaria pre-exposure cohorts, compared to Bagamoyo district population samples from adults 18-45 years (n=122); \**P<0.05*, \*\**P<0.01* Kruskal-Wallis test with Dunn’s multiple comparison test. Dotted line: high and low malaria pre-exposure thresholds defined at the 25% percentile (173.8 AU) and 75% percentile (1650 AU) of all n=122 population test samples. (**B**) Kaplan-Meier plot of time to diagnosis in days for high (n=5) and low (n=6) malaria pre-exposure cohorts; *\*P<0.04* Mantel-Cox test. (**C**) *P. falciparum* parasitaemia measured by qPCR over time for each participant; high (n=5) and low (n=6). Mean of triplicate values within assay plotted. C=day of blood-stage challenge. Dotted line: threshold for initiation of treatment (qPCR >500 parasites/µL). (**D**) Parasitaemia outcomes by pre-exposure status and for all participants combined reported as a proportion. (**E**) Model-based simulation analysis estimating the difference in log_10_-parasitaemia area under the curve (AUC) between pre-exposure groups.

**Table 1.**
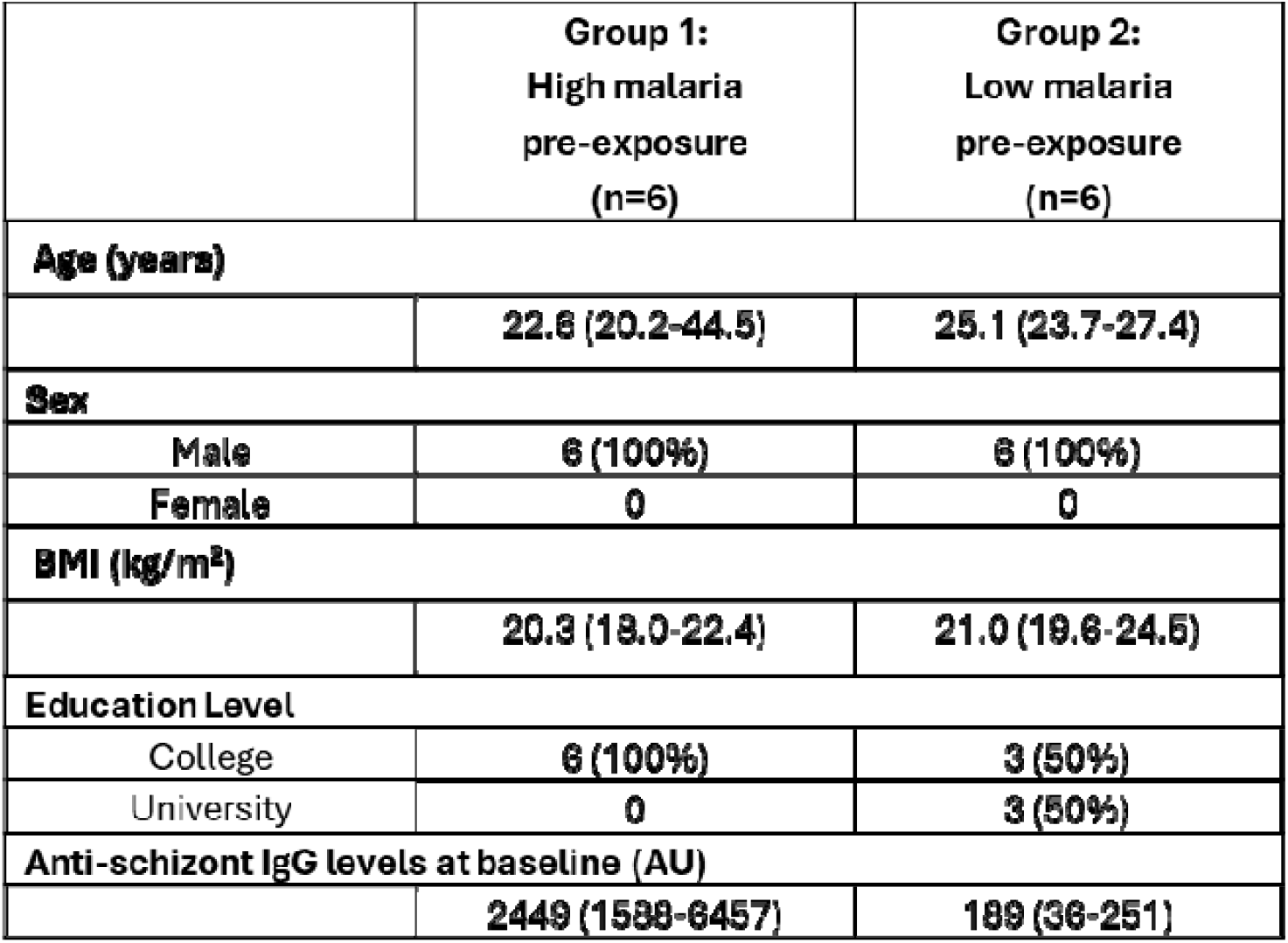
Demographics of VAC083 trial participants and baseline anti-schizont IgG values in each group. Data shown are n (%) or median (range). AU = arbitrary unit by ELISA.

|  | Group 1:<br>High malaria<br>pre-exposure<br>(n=6) | Group 2:<br>Low malaria<br>pre-exposure<br>(n=6) |
| --- | --- | --- |
| **Age (years)** |  |  |
|  | 22.6 (20.2-44.5) | 25.1 (23.7-27.4) |
| **Sex** |  |  |
| Male | 6 (100%) | 6 (100%) |
| Female | 0 | 0 |
| **BMI (kg/m<sup>2</sup>)** |  |  |
|  | 20.3 (18.0-22.4) | 21.0 (19.6-24.5) |
| **Education Level** |  |  |
| College | 6 (100%) | 3 (50%) |
| University | 0 | 3 (50%) |
| **Anti-schizont IgG levels at baseline (AU)** |  |  |
|  | 2449 (1588-6457) | 189 (36-251) |

### Safety outcomes

The cryopreserved stabilate of RBC infected with 3D7 clone *P. falciparum* was thawed on day of challenge (DoC), with the first and last participants inoculated within 2 h 12 min to 2 h 34 min of thaw, respectively. There were no immediate safety concerns post-inoculation for any participant. Two local solicited AEs of mild tenderness at the inoculation site were reported from one participant in each Group (**Supplementary Figure 1**). There were no incidences of pain, pruritus, erythema or inoculation site induration. Post-CHMI solicited AEs differed in frequency between the two Groups with 5/20 (25%) of the total reported solicited AEs occurring in the high malaria pre-exposure group compared to 15/20 (75%) in the low malaria pre-exposure group (**Figure 2A**). In total, five of the six participants in the high malaria pre-exposure group and all six participants in the low malaria pre-exposure group reported at least one solicited AE. Solicited AEs were reported by participants from 1 to 21 days post-CHMI, however, the majority (12/20; 60%) occurred within 24 hours of diagnosis (**Figure 2B**). The most frequently occurring solicited AE was headache, with three events occurring in the high malaria pre-exposure group (two mild, one moderate in severity) and four events occurring in the low malaria pre-exposure group (all mild). The one moderate headache was reported 2.5 days after antimalarial treatment was started and resolved within half a day. Other AEs reported were all mild and included pyrexia, feverishness (subjective fever), rigors, malaise and myalgia in the low malaria pre-exposure group participants; fatigue in two low and one high malaria pre-exposure participants; and back pain in one high malaria pre-exposure participant. All solicited AEs resolved within 48 hours. There were no episodes of chills, nausea or vomiting throughout the period of solicitation for systemic AEs. There were a total of six unsolicited AEs, all mild or moderate and deemed unrelated to the challenge (**Supplementary Table 4**). There were no malaria treatment-associated AEs, no severe AEs, and no serious adverse events (SAEs) during the study.

The most frequent haematological abnormality during the CHMI period was lymphopenia, occurring in five participants (83%) in the low malaria pre-exposure group (three mild and two moderate) and one participant (17%) in the high malaria pre-exposure group (mild) (**Figure 2C** and **Supplementary Table 6**). The minimum lymphocyte counts for the participants in the low malaria pre-exposure group were significantly lower than in the high malaria pre-exposure group (*P*=0.009, Mann-Whitney test), in keeping with decreasing lymphopenia described following repeat CHMI in malaria-naïve participants [2]. There was one case of mild anaemia and one case of mild thrombocytopaenia in the low pre-exposure group and three cases of mild anaemia in the high pre-exposure group. However, there were no significant differences between the minimum haemoglobin or platelet levels in each participant in the two groups (*P*>0.999 and *P*=0.818, respectively, by Mann-Whitney test). All other haematological abnormalities were mild (**Supplementary Table 6**). These abnormal results occurred between 14 and 28 days post-CHMI and all resolved by day 56 post-CHMI.

Clinical biochemistry safety parameters were also largely within normal ranges (**Figure 2C** and **Supplementary Table 6**). The most common biochemical abnormality occurring within 28 days of CHMI was an elevated serum alanine aminotransferase (ALT), as has previously been described in malaria-naïve participants [2]. Three mildly and one moderately raised ALT were observed in participants from the high pre-exposure group. Similarly, two mildly and one moderately raised ALT were seen in the low pre-exposure group. There was no significant difference between the peak ALT levels for each participant in the two groups (*P*=0.937 by Mann-Whitney test). Three cases of hypoglycaemia also occurred in the low pre-exposure group (two moderate and one mild) and one moderate case occurred in the high pre-exposure group. All other abnormal results were mild in severity. All were deemed not clinically significant, and all were within twice the upper limit of normal, with levels returning to within normal ranges by day 56 post-CHMI.

### Parasitaemia outcomes

An *in vitro* viability assay confirmed the blood-stage inoculum contained live parasites that grew in culture (**Supplementary Results**). *In vivo* parasitaemia post-challenge was measured by *Plasmodium* qPCR in all participants (**Figure 3**). In the high pre-exposure group, one participant was retrospectively confirmed to have a wild-type *P. falciparum* infection through genotyping of a C+3 qPCR positive sample and was therefore excluded from all parasitaemia endpoint analyses. This participant remained asymptomatic and was observed in the Clinical Trial Facility up to C+13 when treated. Otherwise, nine of the remaining 11 participants developed parasitaemia detectable by qPCR following blood-stage CHMI between DoC and C+28; 6/6 (100%) and 3/5 (60%) participants from the low and high pre-exposure groups, respectively (**Figure 3B-D**).

Seven of the 11 participants met treatment criteria of either >500 parasites/µL (n=6) or >200 parasites/µL plus malaria symptoms (n=1); 5/6 participants (83%) in the low malaria pre-exposure group and 2/5 participants (40%) in the high malaria pre-exposure group. The remaining four participants did not meet treatment criteria. Two of these, both from the high malaria pre-exposure group, reached C+28 without detectable parasitaemia at any point during the observation period. The remaining two participants, one from each group, had detectable but relatively low levels of parasitaemia throughout the observation period – one in the high malaria pre-exposure group returned to negative parasitaemia on C+28, whilst the other in low malaria pre-exposure group remained parasitaemic at C+28; both were presumptively treated at the end of follow-up on day C+28 (**Figure 3C,D**). Distribution across groups in treatment criteria is shown in **Figure 3D** and **Supplementary Table 8**.

The overall median time to detectable parasitaemia across all 11 participants was 9 days post-DoC, with a shorter median (5 days) in the low pre-exposure group compared to the high pre-exposure group (11 days) (**Figure 3C**). Additionally, time to any pre-defined treatment criteria (i.e. day of diagnosis) was shorter for the low compared to the high pre-exposure group (**Figure 3B**) and clearance was rapid upon treatment for all (**Supplementary Figure 2**). Finally, a post-hoc exploratory analysis observed an estimated difference in area under the curve (AUC) of parasitaemia between low and high malaria pre-exposure groups over days 0-28 post-challenge of 10.6 log_10_(parasites/µL) × days (95% CI: 1.1 – 16.9) (simulation-based two-sided *P=0.030*) (**Figure 3E**).

### Comparison to blood-stage CHMI in malaria-naïve UK adults

We initially performed a study, with n=30 samples of known parasitaemia from a previous UK malaria-naïve adult blood-stage CHMI study [9] in a blinded fashion, to analyse qPCR reporting across sites in the UK and Tanzania and allow for data comparisons. These data showed the UK and Tanzanian qPCR assays were highly correlated but not fully concordant, with a regression slope of 0.9 (**Supplementary Figure 3**). We thus compared the outcomes across these two populations, whilst noting the small discordance between the independent qPCR assays. Following blood-stage CHMI with the same clinical methodology, a higher maximal parasitaemia was reached in the semi-immune malaria-endemic population as compared to the malaria-naïve population, although notably in Tanzania the treatment threshold of 500 parasites per microlitre (p/μL) was 50-fold higher than in the UK where the treatment threshold was 10 p/μL (**Figure 4A**). Median time to detection of parasitaemia by qPCR was four days in malaria-naïve UK adults, as compared to nine days in low malaria pre-exposed and 12 days in high malaria pre-exposed Tanzanian adults; and of the latter group, 2/5 remained negative by qPCR throughout follow up and 1/5 retuned to 0 parasites/μL by C+28 (**Figure 4B,C**).

**Figure 4.**
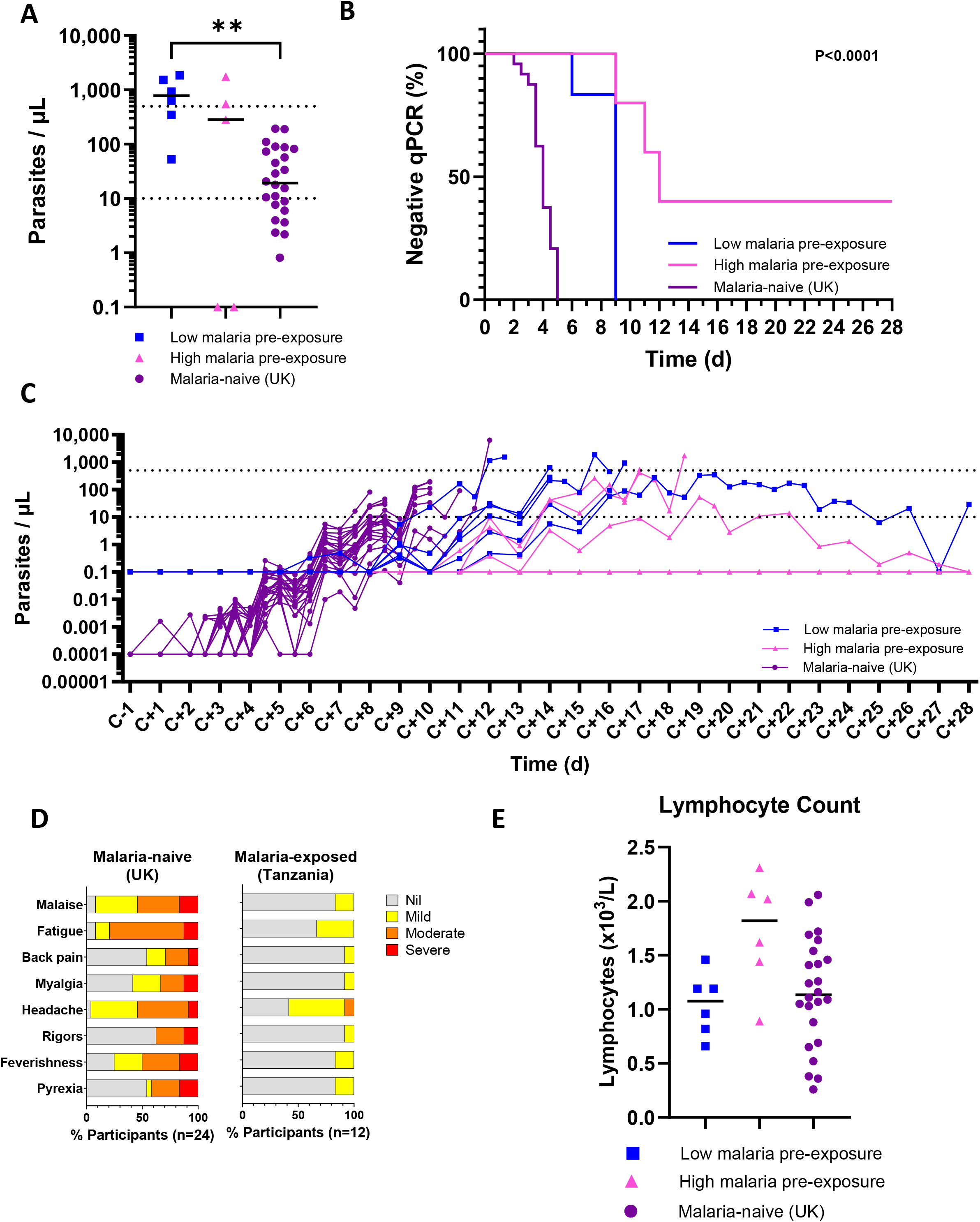
Comparison of outcomes following blood-stage *P. falciparum* CHMI in malaria-naïve UK participants versus semi-immune Tanzanian participants. (**A**) Highest parasitaemia reached for individual participants and median; \*\**P<0.01* Kruskal-Wallis test with Dunn’s multiple comparison test. (**B**) Kaplan-Meier plot showing percentage participants with no detectable parasitaemia by qPCR; *\*\*\*\*P<0.0001* Mantel-Cox test. (**C**) *P. falciparum* parasitaemia measured by qPCR over time for each participant. C=day of blood-stage challenge. Mean of triplicate values within assay plotted. Dotted line: threshold for initiation of treatment (qPCR >10 and >500 parasites/µL for UK and Tanzania, respectively). (**D**) Comparison of solicited AEs reported by malaria-naïve UK CHMI participants (N=24) with those reported by semi-immune Tanzanian participants (N=12). Maximum severity of each AE reported by each participant is shown. (**E**) Comparison of minimum lymphocyte count measured over the CHMI period by participants with high malaria pre-exposure (N=5), low malaria pre-exposure (N=6) and malaria-naïve UK participants (N=24). No significant differences between groups by Kruskal-Wallis test with Dunn’s multiple comparison post-test.

Solicited AEs were reported more frequently, and with higher severity grading, by malaria-naïve participants [2] compared to semi-immune participants despite the higher peak parasitaemia reached by the malaria pre-exposed participants (**Figure 4D**). Comparison of lymphopenia showed a non-significant trend towards a lower minimum lymphocyte count in the low malaria pre-exposure Tanzanian adult group and malaria-naïve UK adult participants [2] as compared to the high malaria pre-exposure Tanzanian adult group (**Figure 4E** and **Supplementary Table 6**).

## DISCUSSION

Here we describe the assessment of the blood-stage CHMI model in a malaria-endemic country using 3D7 clone *P. falciparum*-infected RBC, facilitated by a strong north-south collaboration with mutual sharing of laboratory and clinical expertise. We have demonstrated that blood-stage CHMI was safe and well-tolerated in this cohort of healthy malaria pre-exposed semi-immune male adults, and that clearance of parasites was rapid following treatment.

CHMI studies have been performed in six African countries to-date, incorporating studies of natural immunity and candidate vaccine efficacy [11, 17, 22–32]. These previous CHMI studies have all used cryopreserved sporozoites. The benefits of performing CHMI studies in endemic areas include an increase in the relevance and generalisability of findings to the target population, improved assessment of the interplay between vaccine-induced immunity and naturally-acquired immunity, and the opportunity for capacity building and infrastructure development. Practical challenges can include the availability of *P. falciparum* challenge agents suitable for delivery in endemic settings (stocks are finite and having liquid nitrogen on-site is usually essential), along with local capacity, expertise and infrastructure to run human challenge studies. In addition, undertaking such studies requires building trust with local communities and expertise within local and national ethical committees and national regulatory agencies. Challenges with mosquito-bite CHMI, including potential importation of non-endemic mosquito vector species, have meant that no African country has yet conducted such a study. Here, we report the first blood-stage CHMI study in Tanzania. This involved joint engagement of both the Oxford and Tanzanian research teams with the national regulator, the TMDA, and ethics committee representatives at a very early stage of study conception; this was pivotal to securing timely study approvals and underpinned the success of the project.

Given the anticipated ability of NAI in semi-immune adults to control parasitaemia [33] we split participants into those with relatively low versus high malaria pre-exposure in order to better inform future studies. These groups were defined by anti-schizont IgG serum antibody concentrations as measured in the local population, and are therefore relative to the Bagamoyo area. It would be valuable in the future to apply this approach to other malaria-endemic regions to define relative ELISA thresholds using the same assay and thus help standardise the blood-stage CHMI model between trial sites and countries. Previous reports have suggested alternative stratification by measuring IgG responses to a select panel of *P. falciparum* antigens (including AMA1, MSP1, GLURP and CSP) where participants are classified as seronegative, low, or high exposure based on antigen-specific antibody thresholds derived from reference populations [34]. Epidemiologic variables such as age, site-specific transmission intensity, entomological inoculation rate and reported bed net use have also been considered as proxies for cumulative exposure in malaria vaccine trials [35, 36].

Despite the small group sizes in this study, we did observe significant differences between the low and high malaria pre-exposure groups. A target dose of 1000 infected RBC led to 100% (6/6) infection rate (defined as any detectable parasitaemia) in adults with low malaria pre-exposure. However, only 3/5 (60%) high malaria pre-exposure participants developed detectable parasitaemia, thus giving an 82% (9/11) infection rate across the whole study. 5/6 and 2/5 participants from the low and high malaria pre-exposure groups, respectively, met treatment criteria. Given that reaching pre-defined treatment criteria is often the endpoint in CHMI studies assessing malaria interventions, our data indicate the importance of controlling for prior malaria exposure history between groups in blood-stage CHMI studies in endemic settings. This recommendation is consistent with other CHMI research practice to-date using cryopreserved sporozoites in malaria-endemic regions [23, 24].

Our observations clearly show that a participant infection rate of <100% is likely with this blood-stage CHMI model in an endemic setting and this should be carefully considered when designing control groups for malaria vaccine or drug assessment. Two participants from the high pre-exposure cohort were negative for malaria by qPCR throughout follow-up prior to presumptive treatment on C+28 and one additional participant (also from the high pre-exposure group) returned to negative parasitaemia by C+28 after sustaining a period of low density parasitaemia; given this was the final study day it is unknown whether this participant “cleared” the infection. Notably another participant in the low pre-exposure group also returned to negative parasitaemia on C+27 but bounced back at C+28 when the study ended. The ability of adults to control or clear parasitaemia has been demonstrated following cryopreserved sporozoite challenge [25, 27] but this is the first report of similar observations following blood-stage CHMI. Although we have previously shown no effect of α-thalassemia heterozygosity on parasitaemia following sporozoite challenge in Tanzania [37], investigations to assess for hemoglobinopathies and to characterise the naturally-acquired immune responses that might play a role in control and/or clearance of blood-stage parasitaemia should be undertaken [38, 39]. It is important to consider and control for such variables (and potential modulators of parasitaemia outcomes) when defining future interventional blood-stage CHMI study follow-up periods and endpoints. The possibility of parasite control and clearance also likely necessitates larger sample sizes for blood-stage CHMI trials with semi-immune participants, as compared to equivalent studies in malaria-naïve populations. The detection of a non-3D7 *P. falciparum* infection in one participant on C+3, despite a negative qPCR at baseline, similarly highlights the challenges of conducting CHMI studies with parasitaemia endpoints in endemic areas, even in a dedicated inpatient facility with strict insecticide-treated bed net (ITN) use and indoor spraying, and the importance of accounting for potential participant exclusions from analysis during study design and when performing power calculations [24, 40].

This study had several limitations in addition to the small sample size. Due to lack of existing data in semi-immune populations, we used the same target dose of 1000 infected RBC previously administered to malaria-naïve UK adults [2, 9, 12]. Given our observations here of parasite growth dynamics and the relatively benign symptom profile of infected participants, we would consider increasing the inoculum dose in future studies in malaria-endemic populations as this might improve infection rates in control groups. Additionally, only male participants were included for practical reasons as the study required all participants to be in-patients in one ward for the duration of the challenge period, however, this limits the generalisability of the study findings. Historically, recruitment of females has also been difficult in this particular setting and community engagement is ongoing to address this for future trials. We did not conduct antimalarial drug testing to rule out the presence of antimalarial drugs in the plasma of study participants at screening or post-CHMI, however, the use of antimalarial drugs or drugs with antimalarial activity was deemed unlikely given previous experience with CHMI studies in this population and the close in-patient observation from DoC throughout the in-house observation period.

While we did not adjust the inoculum dose, we pre-defined the treatment thresholds in an attempt to account for the expected impact of NAI. In contrast to blood-stage CHMI studies in the UK, where a treatment threshold of 10 p/µL is routinely used [2, 9, 12, 21], a threshold of 500 p/µL was used in this study as per previous sporozoite CHMI studies in the East African region [23–25]. In endemic regions, the epidemiological definition of a malaria case using the malaria attributable fraction method is typically a fever accompanied by a parasite density ranging from 1,500 to 5,000 p/µL, depending on the local intensity of malaria transmission [41, 42], with a density of 3,000 parasites/µL in adults in a similar transmission setting to Bagamoyo [34]. The chosen threshold of 500 parasites/µL is lower than this, and it is anticipated that the NAI in our participants would offer sufficient protection against clinical disease at this level. Indeed, participants experienced mild, transient laboratory abnormalities similar to those reported in previous endemic studies where experimental malaria infection has been undertaken using sporozoite inoculation [23, 26, 37, 43] and also in blood-stage inoculation studies in non-endemic areas [2, 6, 9, 12, 21]. Noticeably our participants reached 50-fold higher parasite loads with fewer AEs compared to naïve UK adults [12]. Only one participant developed malaria symptoms meeting the treatment threshold – and was then promptly treated and recovered fully. This has been similarly demonstrated in previous CHMI studies in endemic settings [23].

The timepoint for definitive treatment if the treatment threshold has not been reached in UK malaria-naïve CHMI studies is usually 21 days. However, if this timepoint had been used for this pilot study we would have missed observations of parasitaemia returning to undetectable levels, which occurred for two of the participants, between C+21 and C+28, before they received treatment. This highlights the importance for appropriate trial design for the study population. Also related to length of follow-up, in the AUC exploratory analysis, although group-level trajectories were informed by observed data across the full 0–28-day window, later timepoints were supported by relatively few participants, with substantial informative-censoring due to subjects reaching the treatment threshold. Group-level trajectories were thus consequently biased downward at later timepoints, with increasing bias over time and greater in the low-exposed group (low-exposed group 5/6 [83%] censored; median 14 days versus high-exposed group 2/5 [40%]; median 18 days). The AUC difference between pre-exposure groups is therefore biased, potentially conservatively towards the null. These results should thus be interpreted as an exploratory, model-based summary.

The use of blood-stage CHMI as a tool for evaluating blood-stage malaria interventions in endemic settings represents a significant advancement in the effort to accelerate malaria vaccine and drug development. Demonstrating the feasibility of such studies in endemic countries lays the foundation for early-phase trials that can screen potential blood-stage malaria vaccine candidates and antimalarial drug candidates directly in the population that stands to benefit the most from these interventions In conclusion, we have demonstrated that blood-stage CHMI is safe and feasible in semi-immune malaria pre-exposed adults, establishing this model in a malaria-endemic setting of Tanzania for the first time. We have also highlighted the complexities and importance for matched control cohorts to accurately reflect parasitaemia, clearance or lack of detectable infection in malaria intervention reporting. This paves the way for assessment of vaccine and drugs to be tested in target populations to allow for critical down-selection and advancement as well as capacity building across the African continent.

## METHODS

### Trial design and approvals

This study was a single-centre, open-label, blood-stage CHMI trial (called “VAC083”) enrolling adults with varying levels of prior malaria exposure, and thus naturally-acquired immunity, from Bagamoyo district, Tanzania. Specifically, 12 healthy male adults aged 18-45 years were allocated in parallel into two groups of “high” (n=6) and “low” (n=6) malaria pre-exposure, as determined by their baseline serum anti-schizont antibody levels (AU) against defined thresholds. This study was conducted in accordance with Good Clinical Practice. The VAC083 protocol was approved by the institutional review board (IRB) of the Ifakara Health Institute (IHI) (Ref. IHI/IRB/No:06-2022), the Oxford Tropical Research Ethics Committee (ref 6-21), the National Institute for Medical Research Tanzania (NIMR/HQ/R.8a/Vol.IX/4001) and Tanzania Medicines and Medical Devices Authority (Ref. BD.59/62/44/03), and registered at ClinicalTrials.gov (NCT04788862).

### Participants

Participants were graduates or students from higher education institutions who provided written informed consent and underwent screening before enrolment and group allocation. Key exclusion criteria included: confirmed detectable malaria parasitaemia by qPCR on day before challenge; clinically significant comorbidities; hepatitis B, C or HIV seroconversion; and recent use of antimalarial drugs or drugs with antimalarial activity. History of recent antimalarial drug use was assessed via participant report and medical record review. The study exclusively enrolled male participants for practical reasons as the study required all participants to be inpatients in one ward for the duration of the challenge period. A comprehensive list of exclusion and inclusion criteria is provided in the **Supplementary Methods**. The original blood-stage inoculum donor had tested positive for cytomegalovirus (CMV) and Epstein-Barr virus (EBV) [15]. The inoculum has since tested negative for both viruses and there has been no reported seroconversion in >500 participants who have received the inoculum, produced from the same bank of frozen stabilates, in subsequent studies [12]. Therefore, participants were not excluded on the basis of CMV and EBV sero-negativity as had been previously done in early trials [44].

### Blood-stage CHMI

The cryopreserved blood stabilate, containing erythrocytes infected with chloroquine-susceptible *P. falciparum* (3D7 clone), was thawed and prepared under strict aseptic conditions as previously described [2, 9, 12]. Briefly, a single vial containing infected erythrocytes (Queensland Institute of Medical Research Berghofer Master Cell Bank), successfully used to challenge >500 participants in over 40 studies to date, was thawed in a dedicated preparation laboratory, deglycerolised using decreasing saline concentrations with centrifugal wash steps and prepared for injection. The injection volume per participant was 5 mL with a target dose of 1000 parasitised erythrocytes (iRBC) based on estimates of both the donor’s parasite density prior to initial blood banking/cryopreservation [15] and RBC counts upon thawing. An aliquot of the inoculum was assayed to assess parasite viability (further details in **Supplementary Methods**). All 12 participants were inoculated on the same day via an intravenous (IV) cannula within 2h 34min of thaw. This was followed by close inpatient monitoring at the Kingani Clinical Trial Facility at the IHI in Bagamoyo, Tanzania until the successful completion of treatment, confirmation of parasitaemia clearance and clinical assessment by study clinicians.

Once participants met treatment criteria, they received a 3-day regimen of Artemether-Lumefantrine as per label indication (**Supplementary Table 5**) and a single low dose of primaquine (0.25 mg/kg) to clear any gametocytes. Study clinicians could prescribe antipyretics for symptom relief as needed. qPCR was also conducted daily post-treatment initiation until two consecutive negative qPCR results were recorded for each participant prior to discharge from the Clinical Trial Facility.

The Clinical Trial Facility ward was prepared by indoor residual spraying (IRS) prior to the start of the VAC083 study and all participants and clinical staff were equipped with ITNs and mosquito repellents throughout their stay. Study clinical staff were on hand at all times for scheduled clinical assessments and in case of any medical concern. After discharge from inpatient observation, each participant was followed up as an outpatient to day 98 post-challenge (C+98).

### Safety assessment

Participants underwent baseline assessments for vital signs (pulse rate, blood pressure, axillary temperature and respiratory rate), electrocardiography, COVID-19 rapid antigen test and other safety laboratory parameters (full blood count, renal profile, liver function tests and glucose) and were then closely observed for any immediately occurring AEs within the first 2 h following blood-stage malaria inoculum administration. Clinical assessments were conducted twice daily from day of challenge (DoC) until discharge and included vital signs as well as collection of solicited and unsolicited AEs. Haematological and biochemical safety parameters were also evaluated at C+7, C+14, C+21, C+28, C+56 and C+98. The full Schedule of Events is shown in **Supplementary Table 1,** solicited adverse events grading shown in **Supplementary Table 3** and local laboratory abnormality grading scale in **Supplementary Table 7**.

Local AEs at the site of inoculation were collected from DoC to C+2. Systemic AEs were solicited from participants during the period from DoC to C+28 (or until a participant who met malaria treatment criteria [as described below], was treated and discharged from the Clinical Trial Facility). Unsolicited AEs were collected from DoC to C+98. Participants were monitored for malaria treatment-associated AEs from start of treatment until 28 days after the successful completion of treatment. The severity and relationship to the challenge of all reported AEs were assessed by study clinicians. Progress of the trial and safety of participants was also overseen by an independent Safety Monitoring Committee, whilst the safety-related primary objective in the VAC083 study was assessed by the frequency and severity of clinical and laboratory AEs from DoC until end of follow up (C+98). Additional details are provided in **Supplementary Methods** and **Supplementary Tables 3 and 7**.

### Parasitaemia monitoring

Venous blood samples were collected for *P. falciparum* parasite detection and quantification once daily from C+1 which then increased to twice daily sampling during pre-defined morning and evening assessment windows once participants returned positive results with >50 parasites/µL. Parasite detection and quantification were performed using a qPCR assay (PlasQ assay consisting of two independent *Plasmodium* targets combined in a multiplex assay), the Pan-*Plasmodium* 18S rRNA sequence (*Pspp*l8S), and the *P. falciparum* specific var gene acidic terminal sequence (PfvarATS), using the CFX96TM Real-Time PCR Detection System as previously described [45]. The limit of detection for this assay is 0.1 parasites/µL. Additionally, a *P. falciparum* MSP1-, MSP2-DNA based PCR was used to distinguish between the challenge inoculum *P. falciparum* 3D7 clone and the possibility of naturally-acquired *P. falciparum* infection (further details in **Supplementary Methods**) [46]. University of Oxford qPCR methodology for data generation in **Supplementary Figure 3** was performed as previously described [12].

The criteria for diagnosis and initiation of malaria treatment were: a) qPCR result of >500 parasites/μL, irrespective of the presence of symptoms; b) significant signs or symptoms of malaria with a previous qPCR result of >200 parasites/μL. Additionally, participants could be treated on the basis of clinical concern, regardless of the qPCR result. On reaching C+28 without meeting any treatment criteria previously, regardless of qPCR results and/or symptoms, participants were presumptively treated. The >500 parasites/μL treatment threshold has been used previously in malaria semi-immune participants in Kenyan sporozoite-initiated CHMI trials [25] and is below the level of 3,000 parasites/µL reported for symptomatic, clinical malaria in adults in a similar transmission setting [41]. Notably, this threshold is 50-fold higher than the threshold routinely used in malaria-naïve adults in blood-stage CHMI trials using the same inoculum [2, 9, 12]. Pre-defined diagnostic criteria for the data shown in **Figure 4** on N=24 malaria-naïve UK adults under-going blood-stage CHMI have been reported in detail previously [2].

The feasibility-related primary objective in the VAC083 study was assessed by the proportion of participants who developed detectable parasitaemia as measured by qPCR between DoC and C+28, and the proportion of participants who developed sustained parasitaemia detectable by qPCR that spontaneously cleared by C+28. We also report the proportion of participants who met pre-defined treatment criteria during the same follow-up period.

### Viability assay of the blood-stage CHMI inoculum

A viability assay to assess the blood-stage CHMI inoculum was set up on day of challenge in parallel to inoculation of the trial participants. An aliquot of undiluted thawed inoculum was kept at 37°C until the last participant had been challenged, then centrifuged at 800 x*g* for 4 min to pellet the RBC. Supernatant (saline) was removed and replaced with the same volume of complete parasite culture medium. The haematocrit (HCT) level was held at 1%. The iRBCs were then incubated at 37°C for short-term culture with media changes every two days. Seven days later, a thin and thick blood smear of the culture was checked for parasite growth by microscopy following methanol fixation and Field’s stain.

### Anti-schizont ELISA

Serum IgG antibody levels to parasite lysate were assessed by standardised ELISA methodology, as previously described [47–49]. Schizont extract from *P. falciparum* (3D7 clone) parasites produced by the GIA Reference Laboratory, NIAID, NIH, was adsorbed overnight at 4°C to 96 well NUNC-Immuno Maxisorp plates (Thermo Fisher Scientific) at an equivalent of 5×10^2^ parasites per µL. Test sera were diluted in 1% milk in Dulbecco’s phosphate buffered saline (DPBS) and added in triplicate to plates following blocking with 5% milk in DPBS (Sigma). A reference standard and internal control samples from a pool of n=34 high malaria pre-exposed serum samples, from the Bagamoyo Clinical Trial Facility-Ifakara Health Institute (BCTF-IHI) Biobank, plus blank wells, were included. Use of these samples received ethical approval from the Ifakara Health Institute Institutional Review Board (IHI/IRB/No: 49-2020). Bound antibodies were detected using goat anti-human IgG conjugated to alkaline phosphatase (Sigma), developed using 4-nitrophenyl phosphate disodium salt hexahydrate (Sigma) and absorbance at 405 nm (OD405) was determined on a BioTek Elx808 reader with Gen5 software. Antibody units were assigned using the reciprocal dilution of the standard giving an optical density of 1.0 at OD_405_. The standard curve and Gen5 software v3.04 (Agilent) were then used to convert the OD_405_ of test samples to arbitrary units (AU). Low and high malaria pre-exposure thresholds were defined at the 25% percentile (173.8 AU) and 75% percentile (1650 AU) of all n=122 population test samples collected in a serological survey of a study population within Bagamoyo District.

### Statistical analysis

This was a pilot study that aimed to assess the feasibility of blood-stage CHMI in a malaria-endemic setting. The n=12 sample size of the trial was thus designed to only detect very large differences in the incidence of local and systemic adverse events and/or parasitemia endpoints between CHMI groups.

Data were analysed using GraphPad Prism (GraphPad Software, Boston, MA USA). Time to detectable parasitaemia and the treatment threshold was assessed and reported using Kaplan-Meier plots and differences between groups were evaluated using the log-rank test. Immunological and haematological data were analysed using GraphPad Prism (version 10.6.1 for Windows). Kruskal-Wallis tests with Dunn’s correction for multiple comparisons were performed for inter-group comparisons due to small group sizes with a type I error rate set at 0.05. Correlation between qPCR methods was assessed using Spearman’s rank correlation analysis withlinear regression.

As a post-hoc exploratory analysis, a mixed effects model was fit regressing log_10_-transformed PMR measurements on group-specific time trajectories with a shared intercept, with time fit as a third-degree polynomial. A fixed effect for the group-time interaction, and participant-level random effects for the time trajectory were fit. Curves were constrained to a lower limit of –1 on the log_10_ scale to reflect biological plausibility. For each group, the area under the curve (AUC) was obtained by trapezoidal integration of the mixed model’s fixed-effect predictions over days 0–28. Uncertainty was estimated using Monte Carlo simulation, where 10,000 coefficient sets were drawn from the multivariate-normal sampling distribution of the fixed effects and the AUC difference computed for each draw. The corresponding simulation-based 95% confidence interval and two-sided p-value were calculated from quantiles of the resulting distribution. AUC analysis was performed using R version 4.4.3.

## AUTHOR CONTRIBUTIONS

AMM, SJD, AIO and SES conceived the trial.

AMM was the Chief Investigator, AIO was the Site Principal Investigator and SJD was the Senior Laboratory Investigator.

SES, JS, OH, NB, LM, FM, JB, SA, AEK, EK, PK, DQ, OJ, BM, MM, NGM, LMH, MM, BBT, HM, TGM, WFK, IAS, IMM, NSL, FRD, MB, AD, AML, RR, FLN, CAL, SM, MK, JSC, CMN, SJD, AMM,

AIO contributed to the implementation of the study and data collection.

SES, JS, OH, NB, AEK, NGM, NSL, SJD and AMM analysed the data. SA, JSC, FLN and RR performed project management.

SES, JS, OH, CMN, NGM, SJD, AIO and AMM interpreted the data and contributed to writing the manuscript.

All authors had full access to all the data in the study and had final responsibility for the decision to submit for publication.

## Data Availability

All data produced in the present work are contained in the manuscript

## ACKNOWLEDGMENTS

We thank all the VAC083 study participants, all the staff at Kingani Clinical Trials Unit, Ifakara Health Institute, Bagamoyo, Tanzania and the independent Safety Monitoring Committee (Brian Angus, Esther Mwaikambo and Alison Elliott) for overseeing the study. The authors are grateful to Sally Pelling-Deeves for arranging contracts, Susanne Hodgson and Mimi Hou for valuable input in to the study protocol and Jack Quaddy for creating the study database (University of Oxford), James McCarthy for providing the 3D7 inoculum (QIMR Berghofer Medical Research Unit, Australia). This work was funded in part by the European and Developing Countries Clinical Trials Partnership (EDCTP) Multi-Stage Malaria Vaccine Consortium (RIA2016V-1649-MMVC; part of the EDCTP2 programme supported by the European Union); and the National Institute for Health and Care Research (NIHR) Oxford Biomedical Research Centre (BRC). The views expressed are those of the authors and not necessarily those of the National Health Service, the NIHR, or the Department of Health. JS holds a UK Medical Research Council Clinical Research Training Fellowship [MR/Z505134/1].

## FIGURE LEGENDS

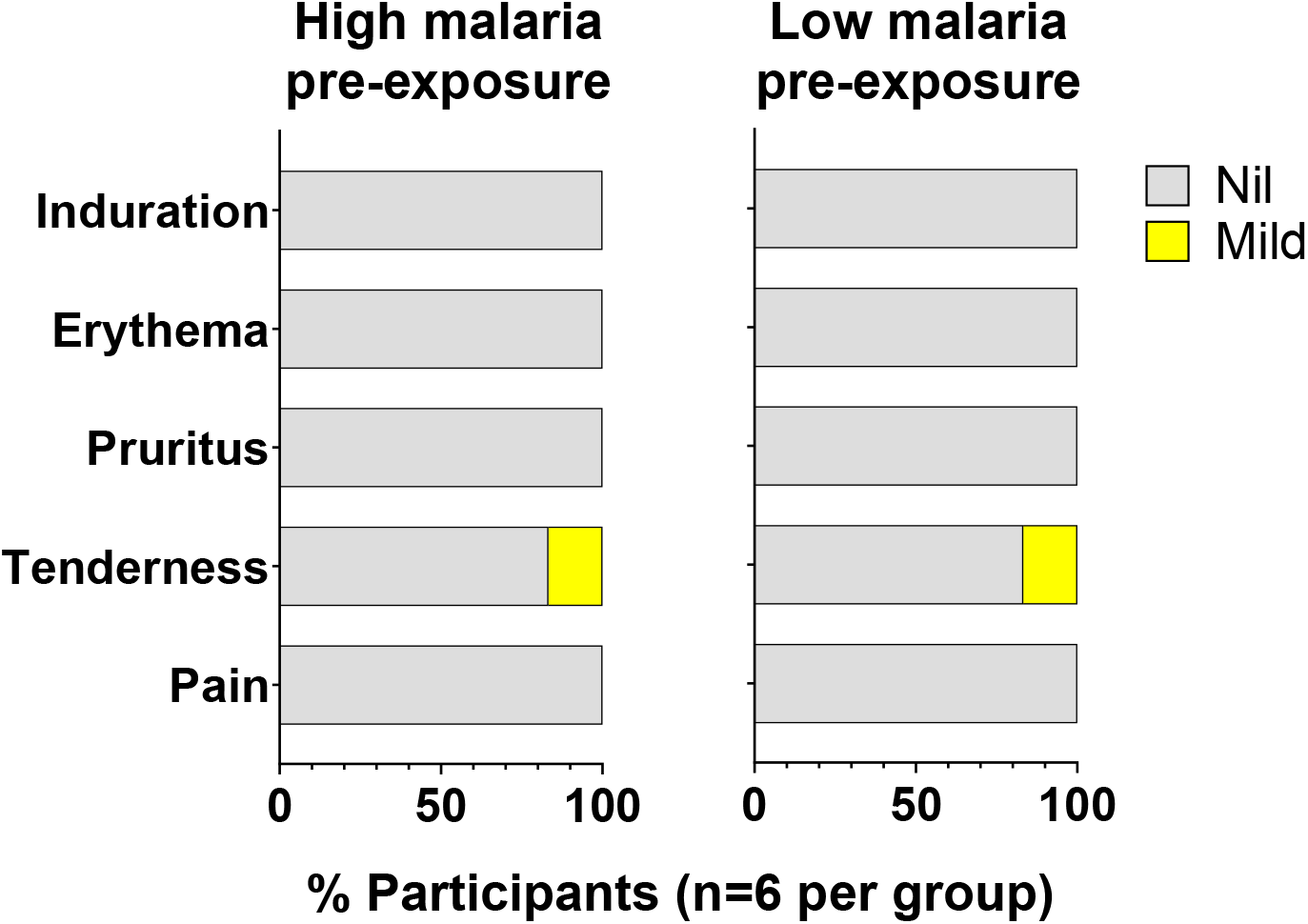
Supplementary Figure 1:

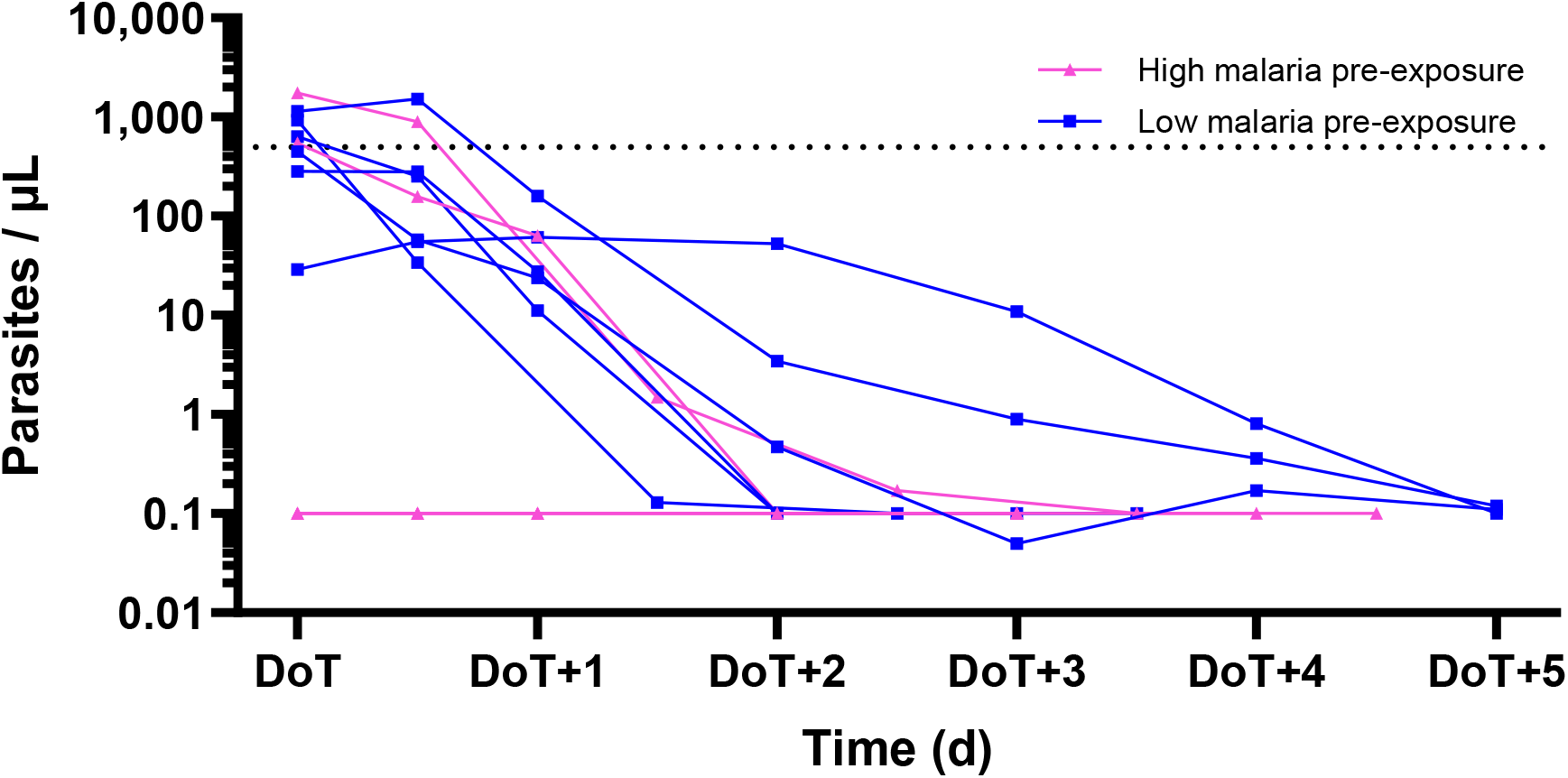
Supplementary Figure 2:

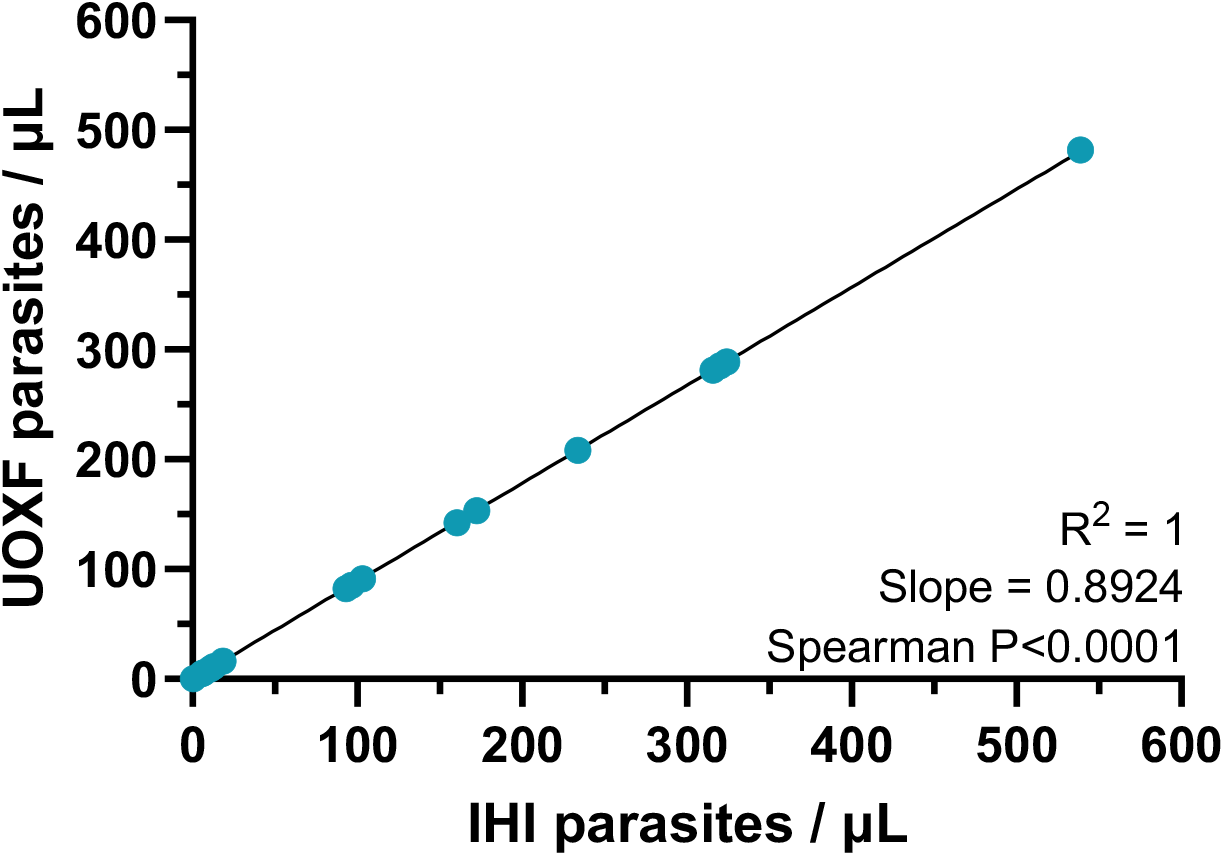
Supplementary Figure 3:

