## Supplementary material for "Establishing a blood-stage controlled human malaria infection model with *Plasmodium falciparum* in healthy semi-immune Tanzanian adults"

### Supplementary Methods

#### Inclusion and Exclusion Criteria for the VAC083 Trial

##### *Inclusion Criteria*

- Participant being adult male aged ≥ 18 and ≤ 45 years, and in good health.
- Participant a resident in Bagamoyo town or rural areas of Bagamoyo district for the past 6 months.
- The participant must be a graduate or currently a student in higher learning institution within the study area.
- Able and willing to complete the informed consent process conducted in Swahili.
- Participant has adequate understanding of the procedures of the study and is able and willing (in the investigator’s opinion) to comply with all study requirements.
- Participant is willing to complete an informed consent questionnaire and is able to answer all questions correctly in a maximum of two attempts.
- Participant is able to communicate well with the investigator and is willing to be monitored in an inpatient setting for 28 days after challenge with infected erythrocytes.
- The participant agrees to refrain from blood donation for 12 months following CHMI.
- Participant agrees to refrain from intensive physical exercise (disproportionate to the participant’s usual daily activity or exercise routine) during the malaria challenge period.
- Participant has signed written informed consent to participate in the trial.

##### *Exclusion Criteria*

- Any history, or evidence at screening, of clinically significant symptoms, physical signs or abnormal laboratory values suggestive of systemic conditions, such as cardiovascular, pulmonary, gastrointestinal, renal, hepatic, neurological, dermatological (e.g. psoriasis, contact dermatitis etc.), allergy, endocrine, malignant, haematological, infectious, immunodeficient, psychiatric and other disorders, which could compromise the health of the participant during the study or interfere with the interpretation of the study results.
- A heightened risk of cardiovascular disease, as determined by: an estimated ten-year risk of fatal cardiovascular disease of ≥5% at screening, as determined by the Systematic Coronary Risk Evaluation (SCORE); history, or evidence at screening, of clinically significant arrhythmia, prolonged QT-interval or other clinically relevant ECG abnormalities; or a positive family history of cardiac events in 1^st^ or 2^nd^ degree relatives <50 years old.
- Body mass index (BMI) of <18 or >30 kg/m^2^.
- A medical history of functional asplenia.
- History of epilepsy in the period of five years prior to study onset, even if no longer on medication.
- Confirmed parasite positive by qPCR a day before challenge i.e., at C-1.
- Screening tests positive for human immunodeficiency virus (HIV), active hepatitis B virus (HBV), hepatitis C virus (HCV).
- Chronic use of i) immunosuppressive drugs, ii) antibiotics, iii) or other immune modifying drugs within three months prior to study onset (inhaled and topical corticosteroids and oral antihistamines exempted) or expected use of such during the study period.
- Any recent or current systemic therapy with an antibiotic or drug with potential antimalarial activity (chloroquine, doxycycline, tetracycline, piperaquine, benzodiazepine, flunarizine, fluoxetine, tetracycline, azithromycin, clindamycin, erythromycin, hydroxychloroquine, etc.) (allowable time frame for use at the Investigator’s discretion).
- History of malignancy of any organ system (other than localised basal cell carcinoma of the skin), treated or untreated, within the past 5 years.
- Any history of treatment for severe psychiatric disease by a psychiatrist in the past year.
- History of drug or alcohol abuse interfering with normal social function in the period of one year prior to study onset.
- Previous participation in any malaria investigational product study (allowable time frame for use at the investigator’s discretion).
- Known hypersensitivity to or contra-indications (including co-medication) for use of chloroquine, artemether-lumefantrine, primaquine or history of severe (allergic) reactions to blood transfusion.
- Participation in any other clinical study in the 30 days prior to the start of the study or during the study period.
- Being an employee or relative of an employee of Ifakara Health Institute.
- Any other condition or situation that would, in the opinion of the investigator, place the participant at an unacceptable risk of injury or render the participant unable to meet the requirements of the protocol.

#### Safety Oversight during the VAC083 Trial

An independent Safety Monitoring Committee (SMC) reviewed the study progress and safety data according to a safety review schedule. A safety report including a list of all reported adverse events and any safety laboratory values outside the normal range was prepared after the completion of in-house observation (C+28) and at the end of the follow-up period (C+98).

#### Malaria Treatment

Following CHMI, all participants diagnosed with malaria were treated with artemether / lumefantrine (ALU) and low single dose primaquine. Artemether 20mg / lumefantrine 120mg was administered orally by a six-dose regimen over 3 days as described in **Supplementary Table 5**. Primaquine was given orally as a single dose of 0.25 mg/kg at the time of diagnosis. Possible side effects related to ALU and primaquine were explained in the informed consent form (ICF). All treatments were administered under direct observation of the clinical investigators. Paracetamol and anti-emetics were available to be used to relieve malaria symptoms such as fever, headache and vomiting.

#### Genotyping of *Plasmodium falciparum* Merozoite Surface Proteins (MSP1 and MSP2)

For DNA extraction the Quick-gDNA Blood Mini Prep Kit from Zymo Research was used. The manufacturer’s instructions were modified as follows:

Whole blood was thawed and vortexed completely, 180 μL transferred into a new 1.5 mL Eppendorf tube. Cells were lysed by adding 720 µL of Genomic Lysis Buffer to 180 µL of whole blood (4:1), vortexed for 5 s and left to stand for 10 min at room temperature. The mixture (total 900 µL) was then transferred to a spin column in a collection tube, centrifuged at 10,000 x*g* for 1 min and eluted.

A 5x PlasQ Primer Mix was prepared using 100 µM stock solutions of all primer and probes.

For the first reaction, 1.0 µL DNA was amplified with 12.5 µL 2x Taq PCR Master Mix, 1 µL forward primer (10 µM), 1 µL reverse primer (10 µM), and sterile ultrapure water 9.5 µL to a final volume of 25 µL.

For the second round of PCR, 1.0 µL first-round PCR products were amplified with a 50 µL reaction system, which included 25 µL 2x Taq PCR Master Mix, 2.0 µL forward primer (10 µM), 2.0 µL reverse primer (10 µM), and water (up to 50 µL).

The primary PCR amplification of the MSP1 and MSP2 genes comprised an initial step of 94°C for 3 min followed by 30 cycles of 94°C for 30 s, 55°C for 30 s, 72°C for 1 min and a final extension of 72°C for 5 min. The nested PCR cycling parameters of the second run are the same as those of the primary reaction. Allelic specific positive control and DNA negative control were included in each set of reactions.

The *P. falciparum* MSP1 and MSP2 genes amplified by nested polymerase chain reaction. PCR products were analysed by electrophoresis on 2.0% agarose gels prepared in 1x TBE (Trisborate EDTA) buffer and stained with 0.01% (v/v) safeview classic gel staining dye . DNA bands were visualised under UV transillumination by a gel document system and fragment sizes estimated by comparison with 100bp DNA ladder from New England Biolabs.

#### Safety Analysis

##### *Adverse Events, Causality and Grading*

###### Adverse Events

Local solicited adverse events after CHMI:

Participants were asked whether they experienced any local adverse events (AEs) from the day of CHMI through to day 2 post-inoculation as listed in the first section of **Supplementary Table 3**.

Solicited systemic AEs:

Participants were asked whether they experienced any systemic AEs from the day of CHMI through to day 28 post-CHMI (or until a participant was diagnosed with malaria, treated and discharged from the clinic) as in **Supplementary Table 3**.

In addition, solicited AEs relating to antimalarial drug treatment were collected during the 3 days of treatment at the facility and followed up for 28 days after completion of treatment via home visits on days 1, 2, 3, 7 and 28 post-treatment.

Unsolicited AEs:

Unsolicited AEs were collected until day 98 after CHMI inoculation as in **Supplementary Table 4**.

##### *Causality*

The causal relationship between the AE and the inoculum injection, malaria infection or study medication was evaluated by the principal investigator (PI). This interpretation was based on the type of event, the relationship of the event to the time of study interventions, and the known biology of the study interventions.

The following guidelines were used to assess the causal relationship:

- No relationship
  - No temporal relationship to the study intervention; and
  - Alternate etiology (clinical state, environmental or other interventions); and
  - Does not follow known pattern of response to the study intervention.
- Unlikely relationship
  - Unlikely temporal relationship the study intervention; and
  - Alternate etiology likely (clinical state, environmental or other interventions); and
  - Does not follow known typical or plausible pattern of response to the study intervention.
- Possible relationship:
  - Reasonable temporal relationship to the study intervention; or
  - Event not readily produced by clinical state, environmental or other interventions; or
  - Similar pattern of response to that seen with similar products.
- Probable relationship:
  - Reasonable temporal relationship to the study intervention; and
  - Event not readily produced by clinical state, environment, or other interventions or
  - Known pattern of response seen with similar products.
- Definite relationship:
  - Reasonable temporal relationship to the study intervention; and
  - Event not readily produced by clinical state, environment, or other interventions; and
  - Known pattern of response seen with similar products.

###### Grading the Severity of Adverse Events

For adverse events other than local swelling, redness/discoloration and pain/limitation of limb movement, for which the severity scales are listed in **Supplementary Table 3**, AEs were graded according to the scale below:

- Grade 1 (mild): awareness of a symptom, but the symptom is easily tolerated and causes no or minimal interference with usual activity.
- Grade 2 (moderate): discomfort enough to cause greater than minimal interference with usual activity.
- Grade 3 (severe): incapacitating; symptoms causing inability to perform usual activities; requires absenteeism or bed rest.
- Grade 4 (potentially life-threatening): symptoms causing inability to perform basic self-care functions OR medical or operative intervention is indicated to prevent permanent impairment, persistent disability or death.

###### Laboratory Adverse Events

Laboratory tests were graded based on the relevant toxicity grading scales relevant to healthy HIV-negative participants with modification to suit local or regional normal values as shown in **Supplementary Table 7**. The abnormal values were categorised as either clinically significant or non-clinically significant based on the study physician’s medical judgement.

#### VAC083 Study Objectives

##### *Primary Objective*

- To assess the safety and feasibility of controlled blood-stage *P. falciparum* human malaria infection in healthy Tanzanian adults with varying prior exposure to *P. falciparum*, through experimental injection of cryopreserved *P. falciparum* infected erythrocytes.

##### *Secondary Objectives*

- To determine the effect of pre-exposure to malaria on parasitemia following controlled blood-stage *P. falciparum* infection.
- To determine if malaria infection following inoculation of *P. falciparum* is caused by the inoculum parasite strain and not wild-type strain.
- To determine the effect of blood-stage CHMI challenge on pre-existing naturally-acquired immunity.

### Supplementary Results

#### Assay of Blood-Stage CHMI Inoculum Viability

A short-term culture of the blood-stage challenge inoculum was set up to assay parasite viability *in vitro*, in parallel to the CHMI. Schizonts, as well as several rings, were detected on day 7 in thin and thick blood smears, indicating that 3D7 clone parasites survived the inoculum preparation and were viable at the time the last participant was challenged.

### Supplementary Tables

#### **Supplementary Table 1: VAC083 Trial Schedule of Events**

Schedule shown for Groups 1+2 from screening out to 98 days post-controlled human malaria infection (CHMI)

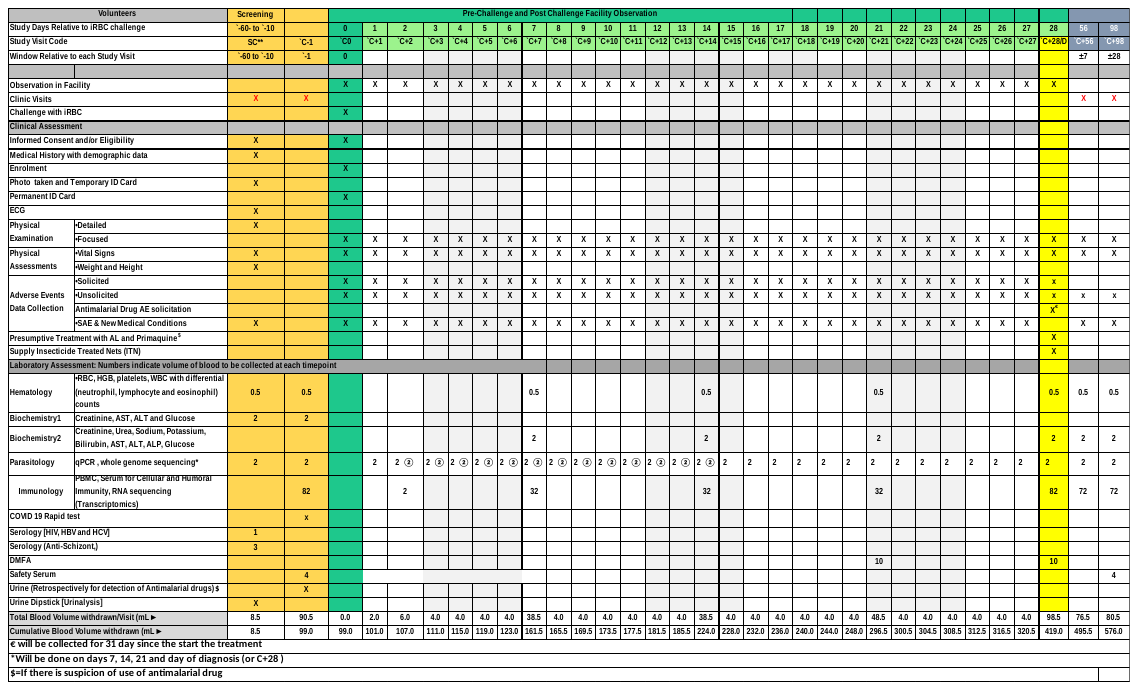

#### **Supplementary Table 2: VAC083 Study Groups**

Group 1 n=6 for high malaria pre-exposure, Group 2 for n=6 low malaria pre-exposure both receiving a planned dose of 1000 infected red blood cells (iRBC)

| **Study Group** | **Prior *P. falciparum* Exposure** | **Number of Participants** | **Planned Dose of iRBC** |
| --- | --- | --- | --- |
| 1 | High | 6 | 1000 |
| 2 | Low | 6 | 1000 |
| Total |  | 12 |  |

#### **Supplementary Table 3: Solicited Adverse Events Grading**

|  |  | Grade | Description |
| --- | --- | --- | --- |
| Local solicited AEs at injection site | ●       Pain  ●       Tenderness  ●       Pruritus | 1 | Daily activity minimally affected, with or without treatment |
|  |  | 2 | Daily activity possible but only with  treatment |
|  |  | 3 | Daily activity not possible even with  treatment |
|  | ●       Erythema  ●       Induration  ●       Bruising  /Extravasated blood | 1 | 2.5 – 5 cm |
|  |  | 2 | 5.1 – 10 cm |
|  |  | 3 | >10 cm, necrosis or exfoliative dermatitis |
| Post-CHMI malaria signs and symptoms | ●       Pyrexia  ●       Feverishness  ●       Chills  ●       Rigors  ●       Sweats  ●       Headache  ●       Nausea  ●       Vomiting  ●       Diarrhoea  ●       Myalgia  ●       Arthralgia  ●       Back pain  ●       Fatigue  ●       Malaise | 1 | Daily activity minimally affected, with or without treatment |
|  |  | 2 | Daily activity possible but only with treatment |
|  |  | 3 | Daily activity not possible even with treatment |
| Malaria treatment associate AEs  (collected until completion of treatment) | ●       Dizziness  ●       Abdominal pain  ●       Palpitations  ●       Itchy skin  ●       Rash  ●       Cough  ●       Insomnia | 1 | Daily activity minimally affected, with or without treatment |
|  |  | 2 | Daily activity possible but only with  treatment |
|  |  | 3 | Daily activity not possible even with  treatment |

#### **Supplementary Table 4: Unsolicited Adverse Events**

Unsolicited AEs were collected for 98 days after CHMI inoculation. All AEs were MedDRA coded and are shown below.

| **MedDRA System Organ Class** | **MedDRA Higher Level Term** | **MedDRA Preferred Term** | **Group** | **Onset (days post-CHMI)** | **Duration (days)** | **Maximum Severity** | **Outcome** |
| --- | --- | --- | --- | --- | --- | --- | --- |
| Respiratory, thoracic and mediastinal disorders | Upper respiratory tract infections NEC | Upper respiratory tract infection | 1 | C+35 | 7 | Moderate | Resolved |
|  |  |  | 2 | C+80 | 10 | Moderate | Resolved |
|  |  |  | 2 | C+43 | 6 | Mild | Resolved |
|  | Nasal congestion and inflammations | Rhinitis allergic | 2 | C+0 | 3 | Mild | Resolved |
| Nervous system disorders | Headaches NEC | Headache | 1 | C+83 | 7 | Moderate | Resolved |
| Skin and subcutaneous tissue disorders | Skin and subcutaneous tissue fungal infections | Fungal skin infection | 1 | C+74 | 21 | Mild | Resolved |

#### **Supplementary Table 5: Artemether Lumefantrine Dose and Schedule**

| **Dose/time** | **Number of tablets** |
| --- | --- |
| 1st dose, at the time of initial diagnosis | 4 tablets |
| 2nd dose, at 8 hours after 1st dose | 4 tablets |
| 3rd dose, at 24 hours after 1st dose | 4 tablets |
| 4th dose, at 36 hours after 1st dose | 4 tablets |
| 5th dose, at 48 hours after 1st dose | 4 tablets |
| 6th dose, at 60 hours after 1st dose | 4 tablets |
| Total number of tablets | 24 tablets |

#### **Supplementary Table 6: Clinical Laboratory Abnormalities**

All participants reporting mild (grade 1) or higher severity laboratory abnormalities in the 28 days after CHMI inoculation are shown. Grade 1 = mild; Grade 2 = moderate; Grade 3 = severe.

| **Laboratory Abnormality** | **High Malaria Exposure (n=6)** | | | | | **Low Malaria Exposure (n=6)** | | | |
| --- | --- | --- | --- | --- | --- | --- | --- | --- | --- |
|  | **No. of Participants** | **Max. Grade** | | | **No. of Participants** | | **Max. Grade** | | |
|  |  | **1** | **2** | **3** |  |  | **1** | **2** | **3** |
| **Leucopoenia** | 2 (33%) | 2 (33%) | 0 | 0 | 2 (33%) | | 2 (33%) | 0 | 0 |
| **Lymphopenia** | 1 (17%) | 1 (17%) | 0 | 0 | 5 (83%) | | 3 (50%) | 2 (33%) | 0 |
| **Neutropenia** | 3 (50%) | 3 (50%) | 0 | 0 | 1 (17%) | | 1 (17%) | 0 | 0 |
| **Anaemia** | 3 (50%) | 3 (50%) | 0 | 0 | 1 (17%) | | 1 (17%) | 0 | 0 |
| **Thrombocytopaenia** | 0 | 0 | 0 | 0 | 1 (17%) | | 1 (17%) | 0 | 0 |
| **Elevated alanine aminotransferase** | 4 (67%) | 3 (50%) | 1 (17%) | 0 | 3 (50%) | | 2 (33%) | 1 (17%) | 0 |
| **Elevated aspartate aminotransferase** | 1 (17%) | 1 (17%) | 0 | 0 | 0 | | 0 | 0 | 0 |
| **Elevated total bilirubin** | 1 (17%) | 1 (17%) | 0 | 0 | 1 (17%) | | 1 (17%) | 0 | 0 |
| **Elevated creatinine** | 0 | 0 | 0 | 0 | 2 (33%) | | 2 (33%) | 0 | 0 |
| **Hyperkalaemia** | 2 (33%) | 2 (33%) | 0 | 0 | 0 | | 0 | 0 | 0 |
| **Hyponatraemia** | 3 (50%) | 3 (50%) | 0 | 0 | 0 | | 0 | 0 | 0 |
| **Hypoglycaemia** | 1 (17%) | 0 | 1 (17%) | 0 | 3 (50%) | | 1 (17%) | 2 (33%) | 0 |

#### **Supplementary Table 7: Local Laboratory Abnormality Grading Scale**

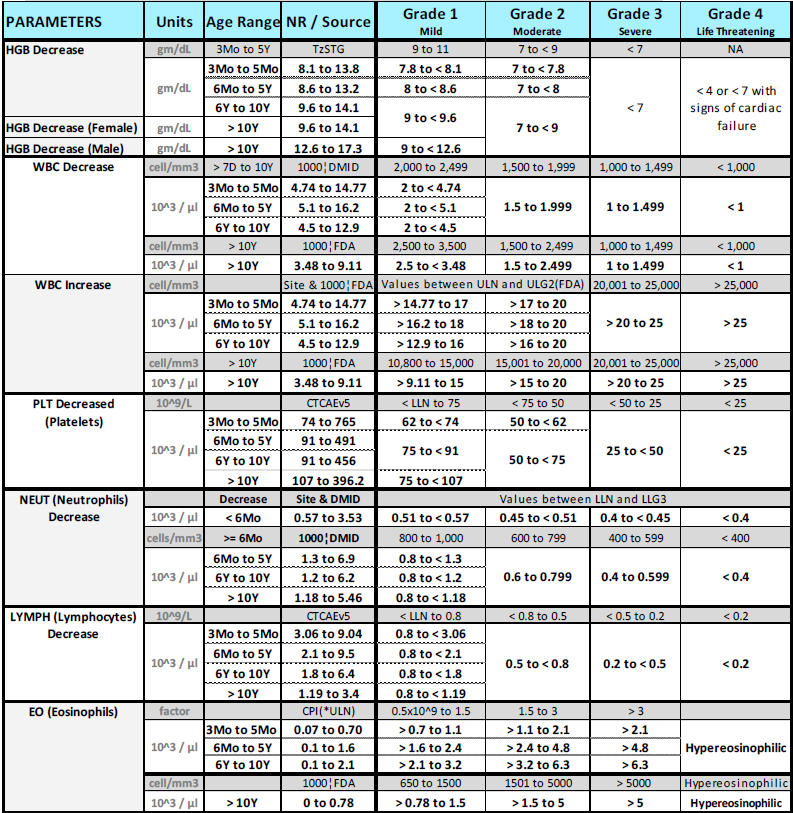

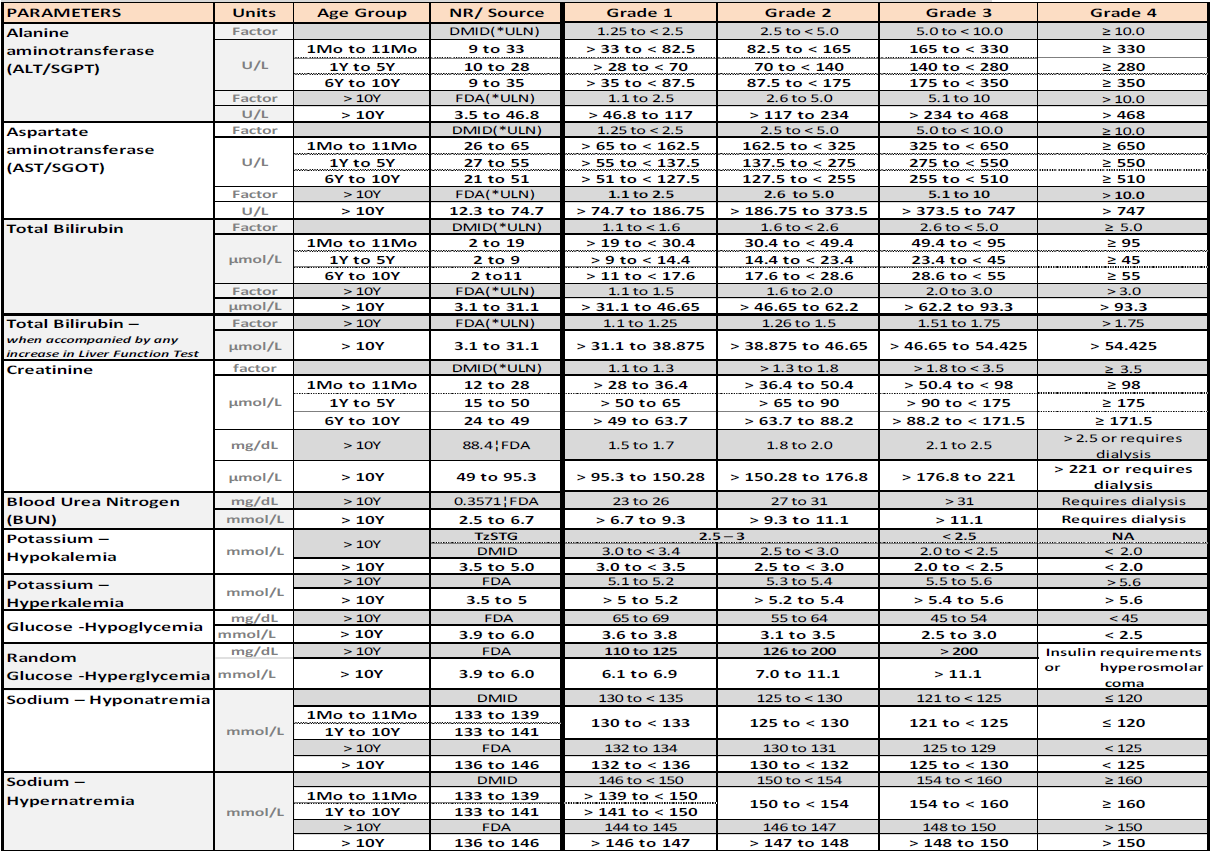

#### **Supplementary Table 8: Parasitaemia and treatment endpoints for participants following CHMI.**

Parasitaemia at the time treatment criteria were reached, the criteria which were met and parasitaemia at the time treatment was initiated per participant.

| **GROUP** | **Parasitaemia/µL at Treatment Criteria or Treatment Initiated Timepoint** | **Treatment Criteria Timepoint** | **Treatment Initiated**  **Timepoint** | **Treatment Endpoint** |
| --- | --- | --- | --- | --- |
| **High** | 0 | C+28 | | C+28 |
| **High** | 547 | C+17 |  | Parasitaemia |
|  | 157 |  | C+17.5 |  |
| **High** | 0 | C+28 | | C+28 |
| **High** | 0 | C+28 | | C+28 |
| **High** | 1746 | C+18.5 | | Parasitaemia |
| **Low** | 1138 | C+12 |  | Parasitaemia |
|  | 1530 |  | C+12.5 |  |
| **Low** | 55 | C+28 | | C+28 |
| **Low** | 1855 | C+15.5 |  | Parasitaemia |
|  | 58 |  | C+16.5 |  |
| **Low** | 932 | C+16.5 | | Parasitaemia |
| **Low** | 636 | C+14 |  | Symptoms |
|  | 252 |  | C+14.5 |  |
| **Low** | 283 | C+14 |  | Symptoms |
|  | 280 |  | C+14.5 |  |

Treatment criteria per protocol:

- Parasitaemia = when the density of infection rises to or past a threshold of 500 parasites per µL.

- Symptoms = if a participant develops symptoms or signs of malaria and a qPCR result within the last 24 h is >200 parasites per µL.

- C+28 = the participant reaches day 28 (C+28) after challenge, without otherwise meeting the above criteria between DoC to C+28.

### Supplementary Figures

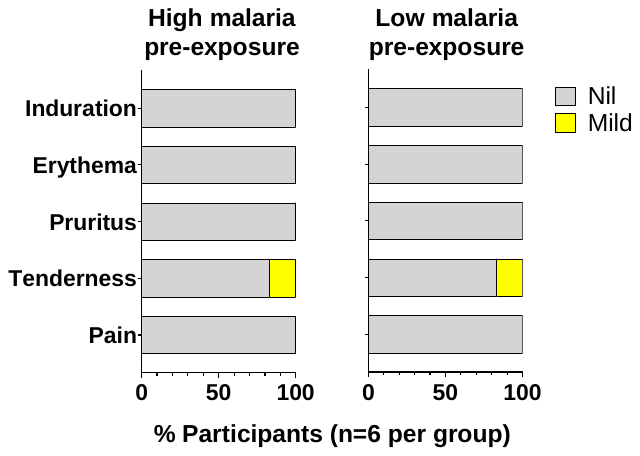

**Supplementary Figure 1.**

**Safety and tolerability of CHMI procedures.** Solicited adverse events (AEs) of induration, erythema, pruritus, tenderness and pain at the CHMI injection site were collected from day of CHMI until two days post-CHMI. The percentage of participants reporting each AE in each group is shown, with the maximum severity of each AE in each participant recorded.

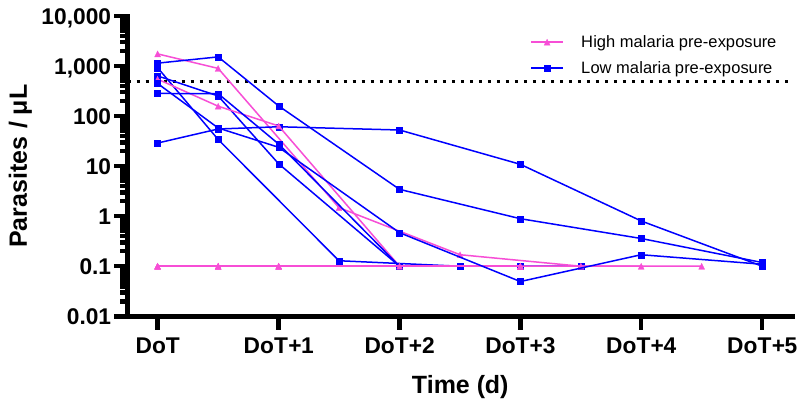

**Supplementary Figure 2.**

**Clearance of parasitaemia.** Parasitaemia as determined by qPCR at Day of Treatment (DoT) and 5 days following treatment (DoT+1-DoT+5). All high (n=5) and low (n=6) malaria pre-exposure participants show reduced parasitaemia following treatment.

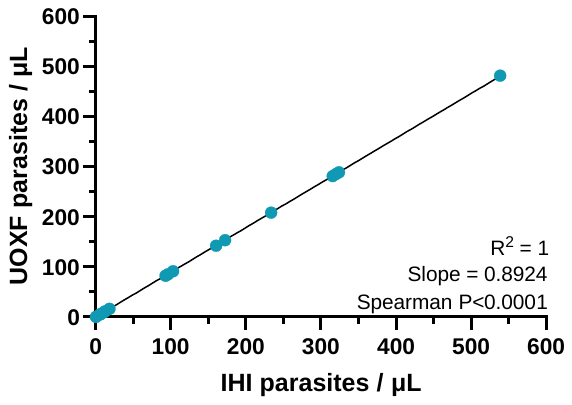

**Supplementary Figure 3.**

**Comparison of qPCR methodologies between blood-stage CHMI study sites.** Parasitaemia as determined by qPCR at University of Oxford, UK (UOXF), and Ifakara Health Institute, Tanzania (IHI), to assess correlation and concordance of assays; Spearman’s *****P<0.0001, R^2^=1. Linear regression slope=0.8924 (95% CI 0.8915 – 0.8934); N=30*
